# A One-Minute Blood Test to Monitor Immune Responses in COVID-19 Patients and Predict Clinical Risks of Developing Moderate to Severe Symptoms

**DOI:** 10.1101/2020.09.30.20203844

**Authors:** Chirajyoti Deb, Allan N. Salinas, Aurea Middleton, Katelyn Kern, Daleen Penoyer, Rahul Borsadia, Charles Hunley, Vijay Mehta, Laura Irastorza, Devendra I. Mehta, Tianyu Zheng, Qun Huo

## Abstract

Coronavirus disease 2019 (COVID-19) has brought enormous loss and interruption to human life and the global economy since the first outbreak reported in China between late 2019 to early 2020, and will likely remain a public health threat in the months and years to come. Upon infection with SARS-CoV-2, the virus that causes COVID-19, most people will develop no or mild symptoms, however, a small percentage of the population will become severely ill, require hospitalization, intensive care, and some succumb to death. The current knowledge of COVID-19 disease progression with worsening symptom complex implicates the critical importance of identifying patients with high clinical risk compared to those who would be at lower risk for disease control and patient management with better therapeutic output. Currently no clinical test is available that can predict risk factors and immune status change at different severity scales. The immune system plays a critical role in the defense against infectious diseases. Extensive research has found that COVID-19 patients with poor clinical outcomes differ significantly in their immune responses to the virus from those who exhibit milder symptoms. We previously developed a nanoparticle-enabled blood test that can detect the humoral immune status change in animals. In this study, we applied this new test to analyze the immune response in relation to disease severity in COVID-19 patients. From the testing of 153 COVID-19 patient samples and 142 negative controls, we detected statistically significant differences between COVID-19 patients with no or mild symptoms from those who developed moderate to severe symptoms. Mechanistic study suggests that these differences are associated with type 1 versus type 2 immune responses. We conclude that this new rapid test could potentially become a valuable clinical tool for COVID-19 patient risk stratification and management.

## Introduction

Coronavirus disease 2019 (COVID-19), an infectious disease that is caused by infection with SAR-CoV-2 virus, has become a global pandemic following an initial outbreak in the People’s Republic of China at the end of 2019 and beginning of 2020. To date, more than 33 million people around the world have been infected with the virus and more than one million people have died of COVID-19 (*1, 2*). Upon infection, while most people will show only mild or no symptoms, a significant portion of the population will develop moderate to severe symptoms that require hospitalization, aggressive treatments such as mechanical ventilation, intensive care, and some eventually succumb to death (*3-5*). The mortality rate based on the number of confirmed positive cases varies from on average 2-3% to as high as ∼10% in some countries (*1*). Symptoms of COVID-19 include high fever, sore throat, dry cough, fatigue, troubled breathing, loss of appetite, body ache, and other health problems (*1, 2*). Pneumonia and extensive lung tissue damages are hallmark pathology in patients who progressed into severe cases and died of COVID-19 (*3-5*). It is likely that COVID-19 will remain as a global public health threat in the months and even years to come.

To control the current pandemic and prepare for future outbreaks, better diagnostic tests are needed not only for the detection of COVID-19, but also for predicting the clinical outcome of the patient. Risk stratification is essential for better management of the patients and health care resources (*6, 7*). Our immune system and function play a critical role in the defense against infectious diseases. While certain types and certain levels of immune response are essential to help the body clear the virus and recover from the infection, an over activated immune response could exacerbate the disease, leading to severe morbidity and mortality (*8, 9*). Since the initial outbreak to present, the study of immune responses in COVID-19 patients has been a central focus of the research and medical community. Extensive studies have found that the immune response in COVID-19 patients is highly complicated, varies significantly from person to person, and patients’ immune response dynamics has a direct association with their clinical outcome (*10-18*). Patients who developed severe symptoms or died of COVID-19 have reduced number of T cells and their immune responses are skewed towards type 2, antibody-mediated immunity (*19-25*). These patients are more likely to experience cytokine storm, production of excessive cytokines that lead to uncontrolled inflammation of the body, tissue damage and eventually organ failure. In contrast, asymptomatic patients or patients with mild symptoms appear to have stronger type 1, cell-mediated immunity than patients with more severe symptoms (*26*). A recent longitudinal study by Lucas et al. revealed additional immune response features that are indicative of divergent disease trajectories and poor clinical outcome in COVID-19 patients (*27*).

Our laboratory has recently developed a rapid blood test, D2Dx immunity test, to monitor the humoral immune status and responses in humans and animals (*28-32*). The test uses a gold nanoparticle (AuNP) as a pseudo virus pathogen to probe the humoral immunity in blood plasma or serum samples, as illustrated in Figure 1. It is widely known that certain nanoparticles will elicit strong immune responses when injected *in vivo* (*33-36*). From our studies, we found that this *in vivo* immune reaction can be replicated *in vitro* in blood plasma or serum samples, and may be used for diagnostic applications. Specifically, our studies found that when the AuNP is mixed with a blood plasma or serum sample, blood proteins, especially the three most important proteins from the immune system, immunoglobulin protein IgG, IgM, and complement proteins, will react with the AuNP as if it is a virus pathogen encountered *in vivo* (*28*). Depending on whether the immune response is type 1 or type 2-biased immunity, these interactions will lead to different degrees of AuNP aggregate formation (*29*). Type 1-biased immune response leads to more extensive AuNP aggregation, and type 2-biased immune response leads to lesser degree of AuNP aggregation (Figure 1A). The reaction product can be detected by measuring the average particle size of the assay solution using a dynamic light scattering technique (*28, 30-32*), or by monitoring the surface plasma resonance absorption change of the AuNPs using a UV-Vis spectrophotometer or colorimeter such as shown in Figure 1B (*29*). Both detection techniques have been well-established and routinely used for detecting AuNP aggregate formation for sensing and bioassay applications (*37-40*). The test involves a single step process, and the results can be obtained in less than one minute. We named this new test as D2Dx (from diameter to diagnostics) because the test is based on AuNP aggregate formation and we initially used particle size measurement for detection (*28, 30-32)*. More recently, we have changed the detection method to a lower cost, and more convenient colorimetry method (*29*).

**Figure 1.**
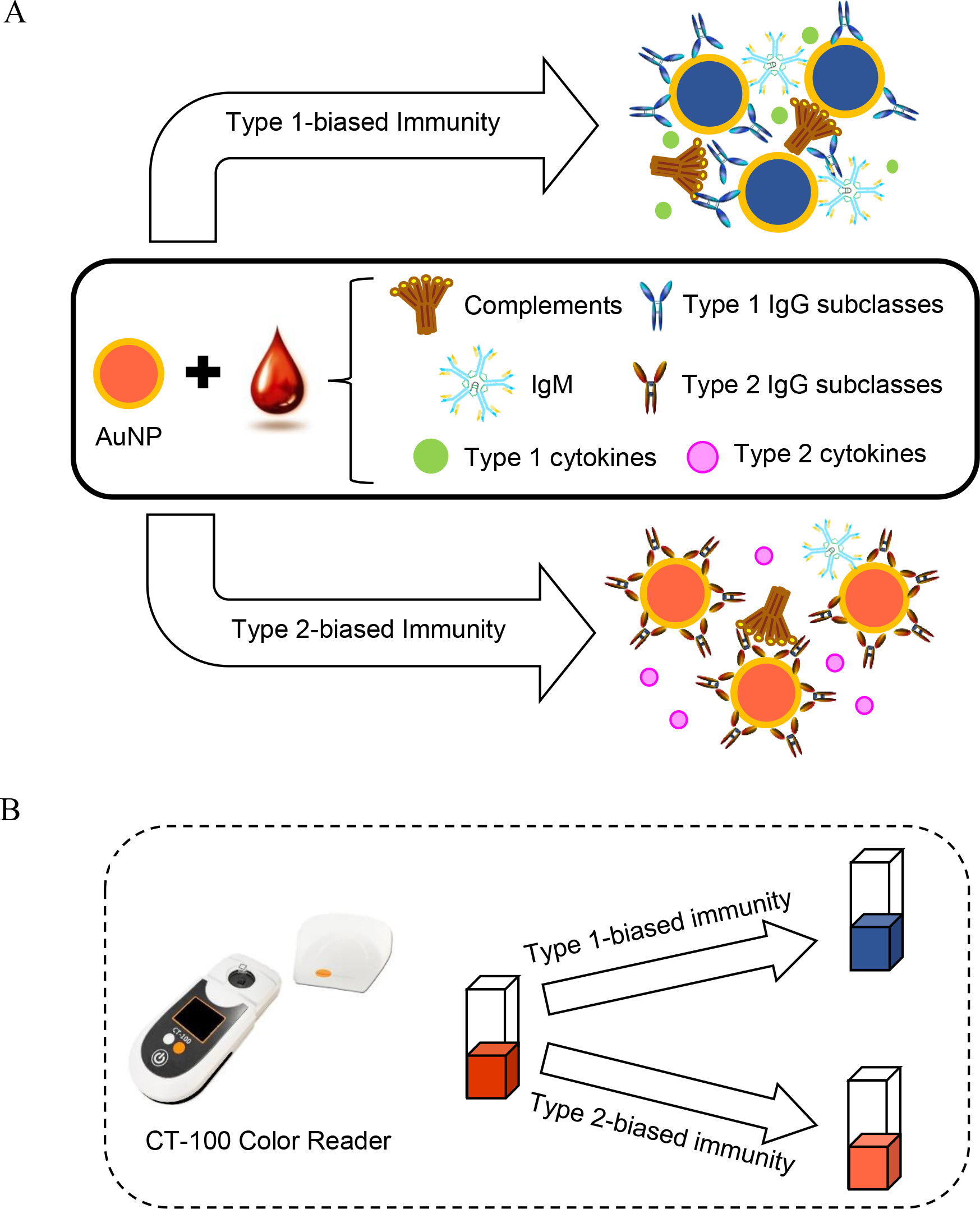
Illustration of the principle of D2Dx immunity test. A gold nanoparticle (AuNP) is used to probe the humoral immune status in a blood plasma or serum sample. The immune interaction between the AuNP and the blood proteins is detected by monitoring the color change of the AuNPs.

In a series of studies we previously reported on laboratory animal models and large agricultural animals, particularly cattle, we have collected extensive experimental evidence that suggests the interaction between AuNPs and blood plasma or serum reflects precisely or very closely the humoral immune status in animals. To give a few examples, we found that the D2Dx immunity test score of mouse models and young calves is correlated directly to the humoral immunity development of the animals from neonates to adults (*28*). We found the D2Dx immunity test score changes dramatically in animals during pregnancy and parturition, and these changes reflect precisely the type of immunity changes that are expected during the reproduction process (*29, 30*). We also observed clearly the test score change in animals with virus infections compared to non-infected controls, and these test score changes mirror almost exactly the humoral immune responses in animals upon infection (*28, 31*).

Based on our understanding on this new test and the large amount of evidence obtained from animal studies, we hypothesize that the D2Dx immunity test may be able to detect the immune responses from COVID-19 patients. In this study, we tested 153 COVID-19 patient samples and 142 negative controls from healthy donors. We detected statistically significant differences between healthy donors and COVID-19 patients; and between COVID-19 patients with different levels of clinical severity. Mechanistic study suggests that the D2Dx immunity test score is associated with the difference in type 1 versus type 2 immunity in COVID-19 patients. Our observation and conclusion are in good agreement with many of the findings made by others on the immune responses to COVID-19. While more extensive validation studies are needed to fully establish the clinical value of this test, we believe the D2Dx immunity test could provide a valuable tool to monitor the immune responses in COVID-19 patients and the test results may be used to predict the risk level and clinical outcome of individuals upon infection with the virus. Because of its simplicity and fast speed, the test may be applied in point-of-care clinical settings. The test may also be used as a research tool to monitor and study the immune responses in COVID-19 patients, provide further guide to vaccine and new therapeutics design and development for COVID-19.

## Materials and Method

### Human Blood Samples

Human blood samples (EDTA K3 plasma) used in this study were obtained from two sources. For healthy controls, we previously obtained and archived 80 blood plasma samples in December 2018 from Boca Biolistics (Boca Raton, Florida) through a different study. The samples were received as de-identified samples. According to NIH policy on human subject research, the use of these samples is considered as non-human subject research, therefore Institutional Research Board (IRB) approval is not needed. Among the 80 samples, 42 were collected in one of the states in the United States and 38 samples were collected in Dominican Republic. All donors were healthy donors without any reported or known infectious diseases when the samples were collected. The samples were aliquoted upon arrival and stored at −80°C until current testing. Prior to testing, the samples were thawed at 4°C overnight, and then left to equilibrate at room temperature for two hours before testing. The sample was tested without dilution or any other treatment.

At Orlando Health, we obtained both COVID-19 positive and negative samples from patients and healthy volunteer donors, 62 from healthy donors and 153 from COVID-19 positive donors. The study (OH IRB # 20.095.06) was approved by Orlando Health IRB#2. Informed consent was obtained from each patient. COVID-19 patients were treated at various hospitals within theOrlando Health hospital system as in-patients or out-patients. The IRB approval also allowed us to use remnant samples from patients and volunteers from a previous serology validation study. The clinical status of the study patients were obtained from the patient’s medical record, or by self-report from volunteers.

### D2Dx immunity test of blood plasma samples

D2Dx immunity test kits (catalog D2Dx-hu-500, lot number hu08012020) were received from Nano Discovery Inc. (Orlando, Florida). Each kit contains the AuNP reagent and cuvettes for 500 tests. A handheld colorimeter reader device, CT-100 from Nano Discovery Inc. was used to read the test result. The specific composition and chemical structure of the AuNP reagent is proprietary information of Nano Discovery Inc. The AuNP reagent was manufactured, formulated and calibrated using the CT-100 reader according to an internal quality control standard established by Nano Discovery Inc.

To perform the test, 50 μL of the AuNP reagent solution was first placed into a cuvette using a micropipette. Then 10 μL of an undiluted blood plasma sample was added. After mixing the assay solution for 5 seconds using a mini-vortex mixer, the cuvette was placed in the cuvette holder in CT-100, and the result was read automatically in 30 seconds. The response of the test was reported directly as the absorbance change of the assay solution over a 30-second of reaction time.

### Kinetic interaction study of AuNP with IgG subclasses

The study of the interaction between the AuNP reagents and IgG subclasses from bovine, human and murine was conducted using the following materials: Bovine IgG1 (pep003, Bio-Rad, 1 mg/mL); bovine IgG2 (pep004, Bio-Rad, 1 mg/mL); human IgG1 (ab90283, Abcam, 3 mg/mL); human IgG2 (ab90284, Abcam, 2.2 mg/mL); human IgG3 (ab118462, Abcam, 2.2 mg/mL); human IgG4 (ab183266, Abcam, 1.5 mg/mL); mouse IgG1 (02-6100, Thermofisher, 1 mg/mL); mouse IgG2a (02-6200, Thermofisher, 1 mg/mL); mouse IgG2b (02-6300, Thermofisher, 1 mg/mL); mouse IgG3 (IMG5119A, Novus Biological, 0.5 mg/mL).

The kinetic study was conducted using a LaMotte model 3250 colorimeter. To an optical cuvette, 100 µL AuNP reagent from the D2Dx immunity test kit (D2Dx-hu-500) was added. Then 10 µL of the IgG subclass protein solution was added. Following mixing for 5 seconds using a mini vortex mixer, the cuvette was placed in the colorimeter, and the absorbance change of the assay solution was recorded every 30 seconds for a total reaction time of 3 minutes.

### Statistical Analysis

Statistical differences of test results between different cohorts were analyzed using student t test, two-sample assuming unequal variances. P values <0.05 were considered as significant difference. The numbers of asterisks indicate significance levels of P values, for example, the symbols of *, **, ***, and **** represent P values of ≤0.05, ≤ 0.01, ≤ 0.001, and ≤0.0001, respectively. If there is no significant difference (P > 0.05) between the groups, the results are presented as “ns”, namely, not significant.

Spearman’s rank-order correlation was used to analyze the correlation between the D2Dx immunity test scores of COVID-19 patients in the severe symptom cohort and the days from symptom onset to blood draw. The strength of the correlation was interpreted according to the scale suggest by Akoglu (*41*): correlation coefficient 1 – perfect; 0.7-0.9 – strong positive correlation; 0.4-0.6 – moderate positive correlation; 0.1-0.3 – weak positive correlation; and 0 – zero correlation. Both student t test and Spearman’s rank-order correlation was conducted using the data analysis function in Microsoft Office 2010 Excel software.

## Results and Discussion

### D2Dx immunity test results of COVID-19 patients versus healthy controls

In this study, we tested 142 negative control samples and 153 COVID-19 positive samples. Table 1 summarizes the clinical status and sample size of each study cohorts. We have three cohorts of negative control samples: Previously, we collected 80 blood plasma samples from healthy donors in December 2018, approximately one year before the COVID-19 outbreak. The samples were collected at two geographic locations: 42 samples from the United States (Normal-USA cohort) and 38 samples from Dominican Republic (Normal-DR cohort). A third cohort of 62 samples were collected at Orlando Health from healthy volunteer donors (Normal-OH cohort) from April and August 2020, the same time period when we collected COVID-19 positive samples. These volunteer donors were tested negative in anti-SARS-CoV-2 IgG and IgM serology test, and never reported any clinical symptoms associated with COVID-19.

**Table 1.**
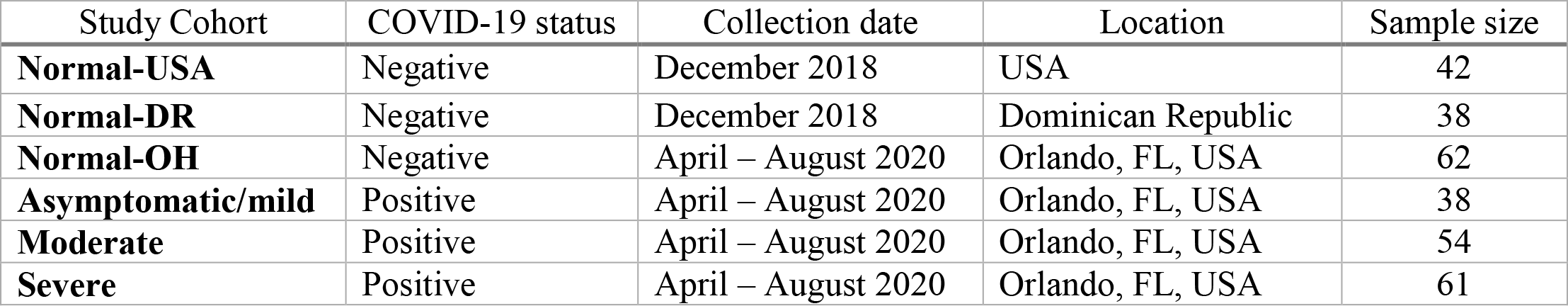
Clinical status, collection dates and sample size of different study cohorts.

| Study Cohort | COVID-19 status | Collection date | Location | Sample size |
| --- | --- | --- | --- | --- |
| **Normal-USA** | Negative | December 2018 | USA | 42 |
| **Normal-DR** | Negative | December 2018 | Dominican Republic | 38 |
| **Normal-OH** | Negative | April – August 2020 | Orlando, FL, USA | 62 |
| **Asymptomatic/mild** | Positive | April – August 2020 | Orlando, FL, USA | 38 |
| **Moderate** | Positive | April – August 2020 | Orlando, FL, USA | 54 |
| **Severe** | Positive | April – August 2020 | Orlando, FL, USA | 61 |

The 153 COVID-19 positive samples were collected between April to August 2020 in Orlando Health (Orlando, Florida). In this study, we used the WHO (World Health Organization) Eight Category Ordinal Scale for Clinical Improvement to rank the clinical severity of the patients and group the patients into different cohorts (*42*). This scale was also used as in the seminal Remdesivir trial (*43*). The uninfected controls are assigned with 0 score. Patients who were tested positive, but exhibited no or only mild symptoms were assigned with a scale 1 (ambulatory with no limitation of activities) or 2 (ambulatory with limitation of activities). These patients were either not hospitalized or were hospitalized for unrelated conditions and found to be positive. From scale 3 and above, all patients were hospitalized and treated in various hospitals at Orlando Health. Blood samples were collected during patients’ stay in the hospital. The documented clinical symptoms were the symptoms exhibited by the patient when the blood sample was collected. According to the clinical symptom severity, the patient was assigned with the Ordinal Scale from 3 – 7: 3 – patients were hospitalized, but no oxygen therapy; 4 – patients require oxygen by mask or nasal prongs; 5 – patients require non-invasive ventilation or high flow mask; 6 – patients require intubation and mechanical ventilation; 7 – patients require ventilation and additional organ support such as vasopressors, RRT, and ECMO. We did not include samples from patients who died of COVID-19 in our study, which would be scale 8. In keeping with the Eight Category Ordinal Scale, patients with scale 1-2 were further grouped as asymptomatic/mild cohort (38 samples); patients with scale 3-4 are grouped as moderate cohort (54 samples); and patients with scale of 5 and above are group as severe cohort (61 samples).

Figure 2 is the D2Dx immunity test scores of the six study cohorts. P values calculated from student t test were listed in the plot for different cohort pairs. Our first statistical analysis was focused on the comparison of the negative control cohort (normal-OH) vs the three COVID-19 patient cohorts, since these samples were collected at approximately the same time period from April to August 2020 from the same clinical site, Orlando Health. Statistically significant difference was observed in the following cohort pairs: normal-OH vs asymptomatic/mild (P value 0.0048); normal-OH vs moderate (P value 1.1E-14); normal-OH vs severe (P value 7E-16), asymptomatic/mild vs moderate (P value 5.25E-06), and asymptomatic/mild vs severe (P value 2.47E-05). The average test score of the normal-OH, asymptomatic/mild, moderate and severe cohort is 0.069, 0.056, 0.029 and 0.032, respectively. The test scores of the moderate and severe cohort are significantly lower than the asymptomatic/mild cohort (P value less than 0.0001).

**Figure 2.**
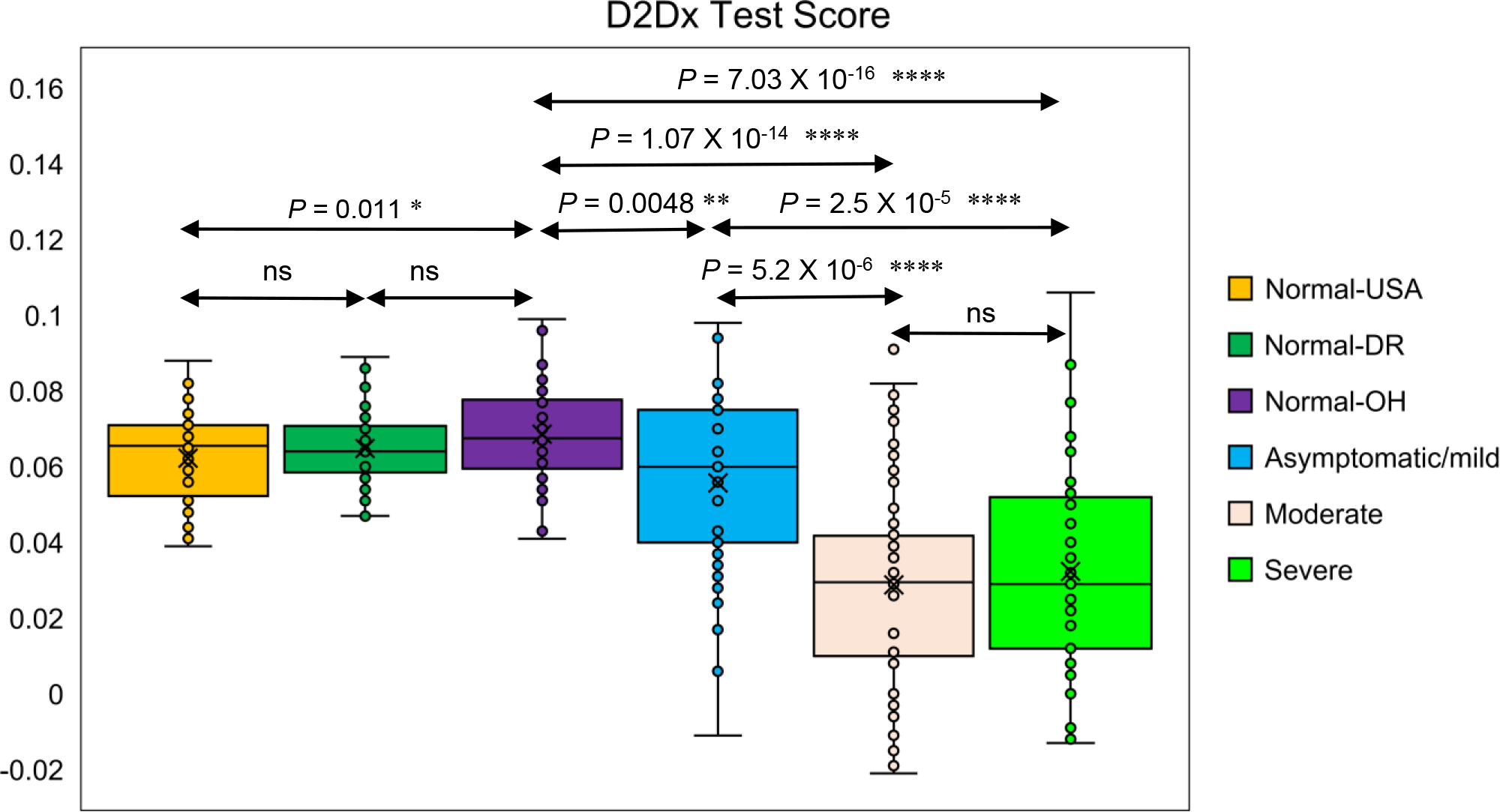
D2Dx immunity test response among various study cohorts.

We then also compared the differences between the three negative control cohorts, normal-OH, normal-USA, and normal-DR. There is no difference between the normal-USA and normal-DR cohort (P value 0.296). Although we observed a slight difference between the normal-OH cohort and the normal-USA or normal-DR cohort (P value 0.01), the average test scores of the three cohorts are very close, in between 0.063 and 0.069. Compared to the difference we observed from the negative control versus the COVID-19 positive samples, this difference is rather insignificant. Furthermore, the samples in the normal-USA and normal-DR cohort have been stored for almost two years before testing. It is most likely that the biological activity of these two cohort samples has changed slightly, leading to a very slightly lower average score in these two cohorts (average score 0.62 and 0.64 for normal-USA and normal-DR cohort) than the more recently collected normal-OH cohort samples (average score 0.69). The comparison of the three negative control cohorts confirms that the D2Dx test is a highly robust test, and the difference we observed from the normal control samples and COVID-19 positive samples are indeed caused by the patients’ active disease status and different immune responses in the patients.

Immune response to viral infection is a highly dynamic process. We analyzed the correlation of the immunity test scores of COVID-19 patients in the moderate and severe cohort with their days from symptom onset and blood collection. This analysis is presented in three graphs: Figure 3A is the correlation analysis of the moderate cohort; Figure 3B is the correlation of the severe cohort by including 21 samples covering symptom onset days from 0 to 27 days; Figure 3C is the correlation of the severe cohort, but including only 16 samples covering symptom onset days from 0 to 14 days. With the moderate cohort, we found no correlation between the symptom onset days and the immunity test scores (correlation coefficient 0.09). With the severe cohort, when all 21 samples are considered, we found a moderate positive correlation (correlation coefficient 0.54) between the immunity test scores and the symptom onset days. When we limit the days from symptom onset to blood collection to 14 days (two weeks) in the severe cohort, a strong positive correlation (correlation coefficient 0.73) was found between the immunity test scores and the symptom onset days. Due to limited data points we have in this analysis, the correlation results should be treated with caution. However, the preliminary data indicates that there appears to be a significant change in the immune response dynamics in COVID-19 patients who develop more severe symptoms, while this change is not seen in patients with moderate symptoms.

**Figure 3.**
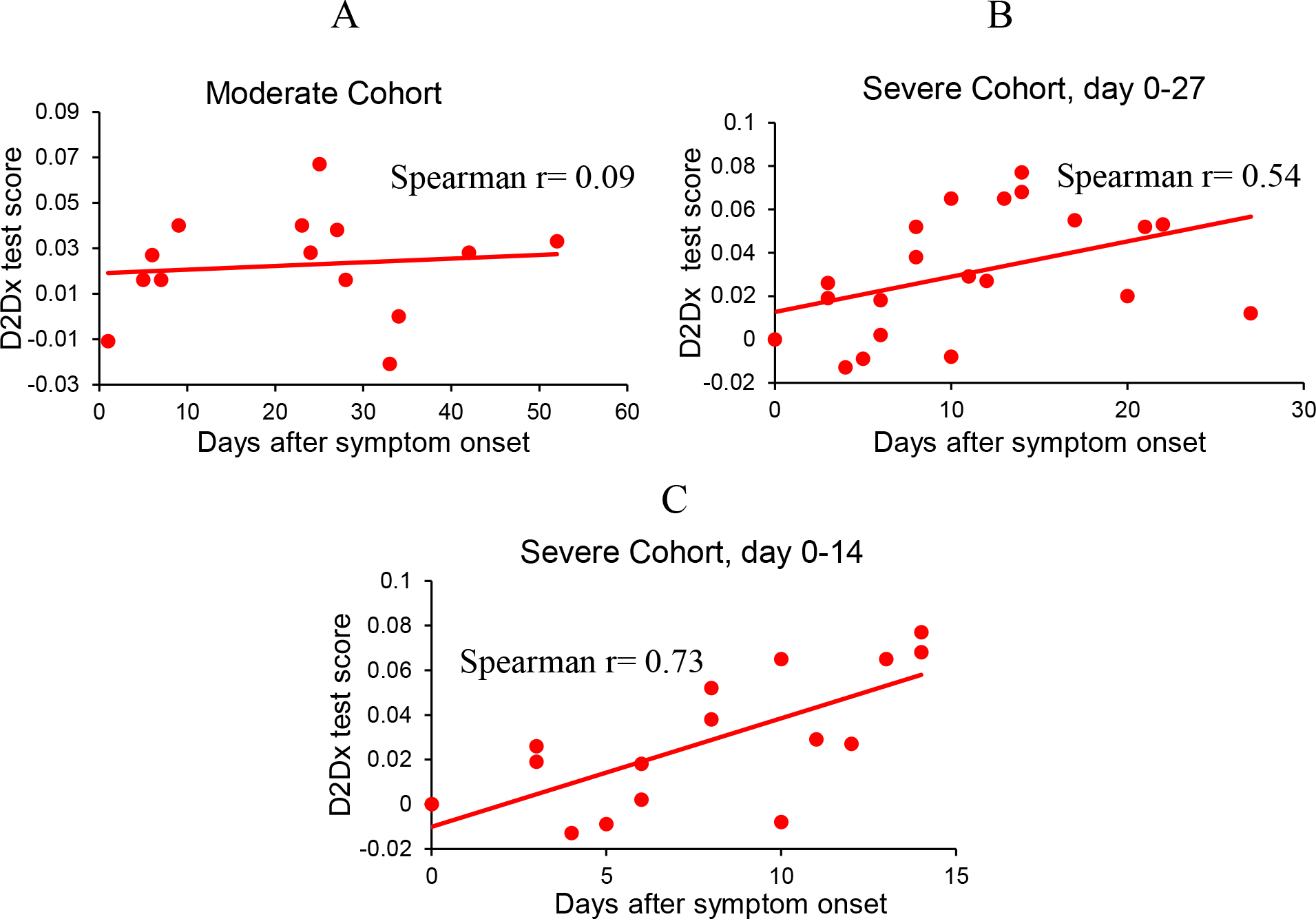
Spearman rank-order correlation of the D2Dx immunity test score of COVID-19 patients in moderate and severe cohort with the days from symptom onset to blood collection. A - Correlation in the moderate cohort (N=14). B - Correlation in the severe cohort with days from the symptom onset to blood draw in the range of 0-27 days (N=21). C - Correlation in the severe cohort with days from the symptom onset to blood draw in the range of 0-14 days (N=16).

### Mechanistic correlation of the D2Dx test score with the immune status of COVID-19 patients

The D2Dx immunity test is not a traditional bioassay that detects a specific biomarker molecule. Instead, the test detects the entire humoral immune response of a blood sample to pseudo virus nanoparticle. A typical humoral immune response involves the coordination and participation of numerous serum proteins such as IgG, IgM, complements, cytokines, chemokines and other biomolecules. Previously, we have shown that the three most important proteins from the humoral immunity, IgG, IgM and complements, are all involved in the interaction with the AuNP probe and impact the D2Dx immunity test score (*28*). Upon infection with SARS-CoV-2, an active immune response will lead to changes in the concentration and distribution of all or some of the immune-related proteins and biomolecules. Hence, it is not surprising that we observed a significant test score change in COVID-19 patients versus normal healthy controls.

In this study, we further demonstrate that the D2Dx immunity test score is reflective of the difference in type 1 versus type 2 immunity in COVID-19 patients. Upon exposure to a virus pathogen, naïve T helper cells (Th0) can polarize into type 1 T helper cells (Th1) or type 2 T helper cells (Th2) (*44-46*). Th1 cells mainly stimulates cell-mediated immune responses, although it also leads to moderate production of certain antibody isotypes and subclasses. Th2-polarized CD4 T cells coordinate the response by eliciting strong antibody-mediated immunity and high antibody titer. Type 1 responses, characterized by intense phagocytic activity, are the default immune response to intracellular pathogens such as viruses (*44*). Type 2 responses, on the other hand, are mounted when type 1 response fails to clear the virus and the infection. Type 2 response can sometime exacerbate the disease and is often associated with severe symptoms and poor clinical outcome. There is extensive evidence suggesting that COVID-19 patients with no or milder symptoms have stronger type 1 immune response, and patients with more severe symptoms tend to exhibit type 2 immune response (*19-26*). There is also evidence that patients with moderate symptoms and type 2 immune response can transition into a type 3 hypersensitivity (*47*), characterized by cytokine storm and severe inflammation that can eventually lead to death. Type 1 and type 2 immunity are inversely modulated by each other: certain cytokines produced by Th1 cells can inhibit the activity of Th2 cells, and vice versa (*44*).

Although the D2Dx test, a test conducted on blood plasma or serum samples, only detects humoral immunity and immune response (because cellular components of the blood are not present), the result can reveal information on cell-mediated immune status as well, especially the balance of the two. This connection is made through the different types of immunoglobulin G protein, IgG subclasses. In humans, IgG1 and IgG3 are associated with type 1 response (cell-mediated response) while IgG4 is linked to type 2 response (humoral response) (*48, 49*). In animals, bovine IgG2 is associated with type 1 and IgG1 is associated with type 2 immunity (*50*). In murine model, IgG2a is a specific indicator of Th1 lymphocytes and mouse IgG1 is a marker of Th2 lymphocytes (*51*).

In our study, we examined the interaction of the AuNP reagent used in the D2Dx immunity test with the different types of IgG subclasses. We analyzed all IgG subclasses from human, bovine and murine. The test was conducted using the same protocol as used for blood plasma analysis. Figure 4A is the reaction kinetics between the AuNP reagent with IgG subclasses indicative of type 1 immunity, i.e., human IgG1 and IgG3, bovine IgG2, and mouse IgG2a. Figure 4B is the reaction kinetics between the AuNP reagent and type 2 indicating subclasses, namely, bovine IgG1, human IgG4, mouse IgG1. The association of human IgG2, mouse IgG2b and mouse IgG3 in type 1 versus type 2 immunity is not as clearly understood as other IgG subclasses, but the results are presented here as well.

**Figure 4.**
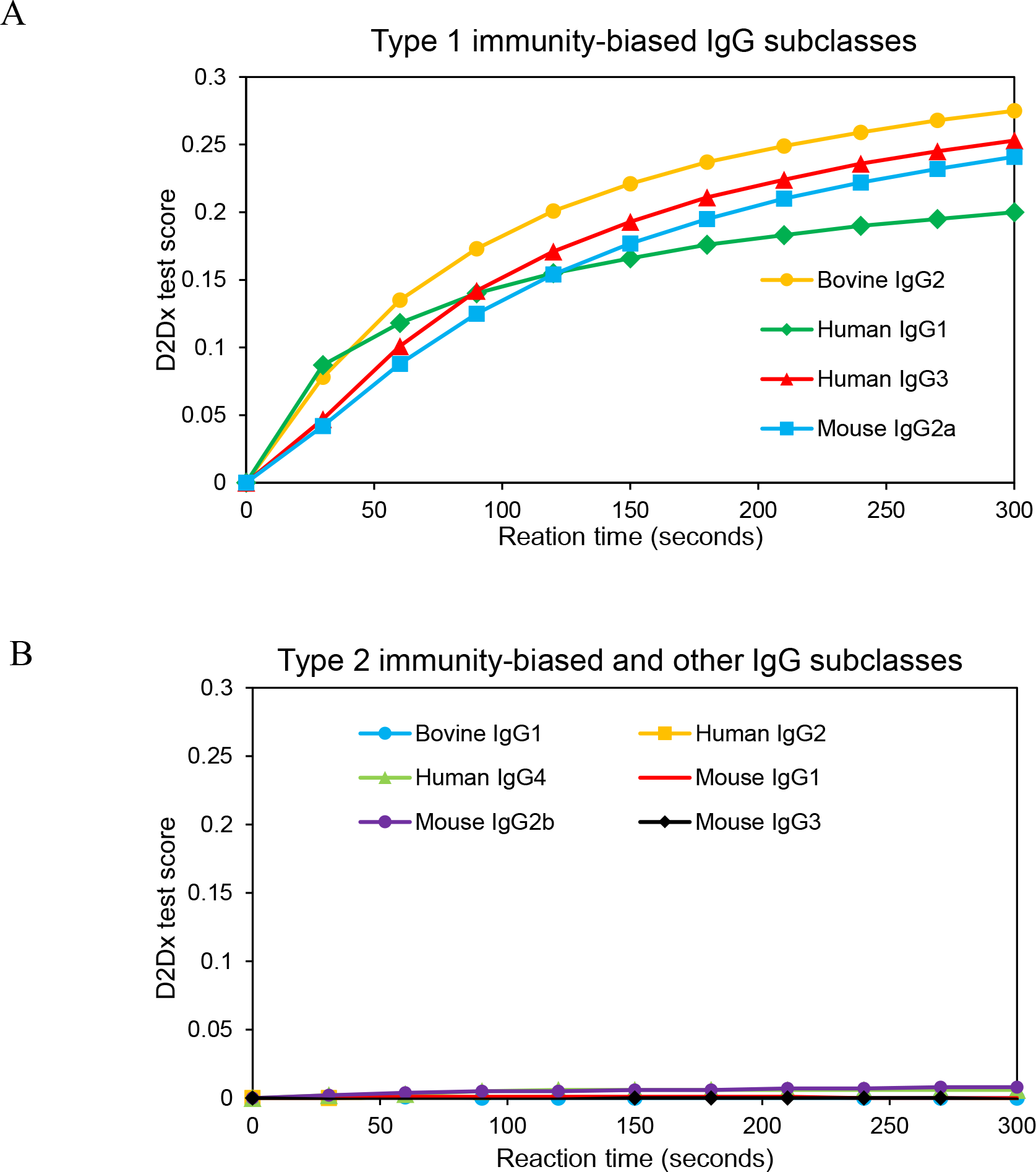
A - Kinetic interaction of AuNP with IgG subclasses indicative of type 1 immunity, including bovine IgG2, human IgG1, human IgG3, and mouse IgG2a. B-Kinetic interaction of AuNP with type 2 immunity – related and other IgG subclasses, including bovine IgG1, human IgG2, human IgG4, mouse IgG1, mouse IgG2b, and mouse IgG3. Kinetic curves shown here are representative of multiple measurements. Graph A and B are presented at the same scale for direct comparison.

Comparison of the two groups of IgG subclasses revealed significant difference between type 1 and type 2 immunity-related subclasses. All type 1-related IgG subclasses, regardless if it is from human, bovine or murine sources, caused dramatic color change of the assay solution; while all IgG subclasses associated with type 2 immunity, led to almost no color change of the assay solution. Translating this finding into blood samples, we can interpret the D2Dx immunity test score of blood samples as the following: a high test score suggests a strong type 1 immunity and weak type 2 immunity; while a low test score indicates a weak type 1 immunity and strong type 2 immunity.

This interpretation agrees perfectly with what has been found regarding the type 1 versus type 2 immunity in COVID-19 patients, and their connection with the clinical severity of the patients. Patients who were tested positive but developed no or mild symptoms must have had strong type 1 immunity. The type 1 immune response is enough to clear the virus and the infection. A Type 2 immune response may be activated in these asymptomatic/mild patients, but most likely to a much lesser degree. Patients who develop moderate to severe symptoms are those who did not or could not mount a sufficient type 1 immune response, leading to an invoked type 2 response to help control the infection. A study by Roncati et al. suggested that there may be a transition from type 2 response to type 3 hypersensitivity in patients who advance from moderate to severe symptoms (*47*). Our limited correlation analysis between D2Dx immunity test scores and days from symptom onset to blood draw in patient cohort with severe symptoms (Figure 3) appears to support this transition theory. The test scores of both severe and moderate cohorts are very low in comparison to normal controls at early stage of their disease, but the scores of the severe cohort increases continuously over time (correlation coefficient 0.73) while the moderate cohort maintains a low score throughout the course. The potential connection between the D2Dx immunity test score and the transition of COVID-19 patients from moderate to severe cases needs to be explored and confirmed in more extensive studies.

## Conclusion

We reported here an extremely simple and rapid test that can detect the immune status change in COVID-19 patients and the differences between COVID-19 patients with milder symptoms versus those with more severe symptoms and poor clinical outcome. The test can be potentially used as a point-of-care clinical test for COVID-19 patient risk stratification and management. The test involves a simple one-step process and the result is obtained in less than 1 minute. Although we used archived blood plasma samples collected through full blood draw in the current study, the test can use blood samples obtained from finger prick, since only 10 µL plasma sample is needed to perform the test. In addition to potential clinical applications, the novel D2DX testing methodology may provide an important tool to study the immunology and immune responses associated with COVID-19, such as vaccine design and development.

## Supporting information

Conflict of Interest Disclosure

## Data Availability

The datasets during and/or analyzed during the current study are available from the corresponding author on reasonable request.

## Acknowledgement

This study is supported by intramural funding from Nano Discovery Inc. and Orlando Health.

## Notes

### Competing Interest Statement

Dr. Huo reports other from Nano Discovery Inc., outside the submitted work; In addition, Dr. Huo has a patent Detection of Interaction Between an Assay Substance and Blood or Blood Components for Immune Status Evaluation and Immune Related Disease Detection and Diagnosis pending.

### Author Declarations

The study (OH IRB # 20.095.06) was approved by Orlando Health IRB#2.

